# Unpacking the behavioural components and delivery features of early childhood obesity prevention interventions in the TOPCHILD Collaboration: a systematic review and intervention coding protocol

**DOI:** 10.1101/2020.12.17.20248435

**Authors:** Brittany J Johnson, Kylie E Hunter, Rebecca K Golley, Paul Chadwick, Angie Barba, Mason Aberoumand, Sol Libesman, Lisa Askie, Rachael W Taylor, Kristy P Robledo, Seema Mihrshahi, Denise A O’Connor, Alison J Hayes, Luke Wolfenden, Charles T Wood, Louise A Baur, Chris Rissel, Lukas P Staub, Sarah Taki, Wendy Smith, Michelle Sue-See, Ian C Marschner, David Espinoza, Jessica L Thomson, Junilla K Larsen, Vera Verbestel, Cathleen Odar Stough, Sarah-Jeanne Salvy, Sharleen L O’Reilly, Levie T Karssen, Finn E Rasmussen, Mary Jo Messito, Rachel S Gross, Maria Bryant, Ian M Paul, Li Ming Wen, Kylie D Hesketh, Carolina González Acero, Karen Campbell, Nina Cecilie øverby, Ana M Linares, Heather M Wasser, Kaumudi J Joshipura, Cristina Palacios, Claudio Maffeis, Amanda L Thompson, Ata Ghaderi, Rajalakshmi Lakshman, Jinan C Banna, Emily Oken, Maribel Campos Rivera, Ana B Perez-Exposito, Barry J Taylor, Jennifer S Savage, Margrethe Røed, Michael Goran, Kayla de la Haye, Stephanie Anzman-Frasca, Anna Lene Seidler, on behalf of the Transforming Obesity Prevention for CHILDren (TOPCHILD) Collaboration

**Author notes:** **Corresponding author** Brittany J Johnson, Caring Futures Institute, College of Nursing and Health Sciences, Flinders University, GPO Box 2100, Adelaide SA 5001.

## Abstract

**Introduction:** Little is known about how early (e.g., commencing antenatally or in the first 12 months after birth) obesity prevention interventions seek to change behaviour and which components are or are not effective. This study aims to 1) characterise early obesity prevention interventions in terms of target behaviours, delivery features, and behaviour change techniques (BCTs), 2) explore similarities and differences in BCTs used to target behaviours, and 3) explore effectiveness of intervention components in preventing childhood obesity.

**Methods and analysis:** Annual comprehensive systematic searches will be performed in Epub Ahead of Print/MEDLINE, Embase, Cochrane (CENTRAL), CINAHL, PsycINFO, as well as clinical trial registries. Eligible randomised controlled trials of behavioural interventions to prevent childhood obesity commencing antenatally or in the first year after birth will be invited to join the TOPCHILD Collaboration. Standard ontologies will be used to code target behaviours, delivery features and BCTs in both published and unpublished intervention materials provided by trialists. Narrative syntheses will be performed to summarise intervention components and compare applied BCTs by types of target behaviours. Exploratory analyses will be undertaken to assess effectiveness of intervention components.

**Ethics and dissemination:** The study has been approved by The University of Sydney Human Research Ethics Committee (project no. 2020/273) and Flinders University Social and Behavioural Research Ethics Committee (project no. HREC CIA2133-1). The study’s findings will be disseminated through peer-reviewed publications, conference presentations, and targeted communication with key stakeholders.

**Discussion:** Our study will provide an in depth understanding of behavioural components and delivery features used in obesity prevention interventions starting antenatally or in the first 12 months after birth. Understanding common intervention approaches in a systematic way will provide much needed insight to advance the design of early obesity prevention interventions and provide the opportunity to undertake future quantitative predictive modelling.

**Registration:** PROSPERO registration no. CRD42020177408

**STRENGTHS AND LIMITATIONS OF THIS STUDY:**

- This study provides an understanding of behaviours targeted, behaviour change techniques and delivery features used in early childhood obesity prevention trials identified in a systematic review as being eligible for inclusion in the Transforming Obesity Prevention in CHILDren (TOPCHILD) Collaboration.
- Extends previous methods by coding behaviour change techniques in published and unpublished intervention materials and performing cross validation with trialists through the TOPCHILD Collaboration.
- Using standardised coding taxonomies will allow for comparisons across studies, and we will pilot test new ontologies from the Human Behaviour Change Project.
- Explores the complex area of targeting parent and caregivers’ behaviours to impact child outcomes across four key obesity prevention behavioural domains (relating to infant feeding practices, food provision and parent feeding practices, movement practices, sleep health practices).
- This study will provide preliminary results regarding the examination of intervention components’ effectiveness based on exploratory analysis. Yet, the internationally unique database this project creates will further our understanding of effective intervention components in future research.
- To date we already have 38 out of 65 eligible trials agreeing to share data, since not all trials may provide unpublished material we may perform sensitivity analyses comparing trials that have shared data to trials that have not shared materials.

## INTRODUCTION

Childhood overweight and obesity are a global concern, with 2019 estimates indicating that 38 million children under the age of five years are affected.^1^ Increasing rates of overweight and obesity have been observed in young children in low- and middle-income countries, highlighting this widespread issue and the overlap of undernutrition and obesity as a double burden for public health systems.^2 3^ The causes of childhood obesity are multifaceted, including genetic, epigenetic, environmental, social and behavioural factors.^4^ Many suggest that obesity prevention should start early in life, if not prenatally or prior to conception, to establish healthy behavioural patterns in young children and avoid metabolic programming that will continue across the life course.^5 6^

Parents and caregivers play a key role in shaping children’s developmental environment and behaviours, particularly in the first year after birth when children are dependent on their parents’ and caregivers’ guidance.^7 8^ While infant behavioural outcomes are the focus for early obesity prevention, parents and caregivers are the key agents of change.^8^ Parents and caregivers should be supported to obtain the knowledge and acquired behaviour to act in ways that provide infants with home environments to develop optimal energy-balance related behaviours, resulting in favourable infant feeding, dietary intake, physical activity, sedentary behaviour, and sleep.

Trials commencing antenatally or in the first 12 months after birth are from here on referred to as early obesity prevention interventions. Many of the first of such complex trials began in 2006-2009 (e.g. ^9-15^). These trials aimed to modify several parent behaviours known to be associated with infant obesity risk. Since the first trials, the number of early obesity prevention interventions has grown substantially, providing an extensive evidence base to inform how we seek to prevent the global issue of childhood obesity. This evidence base continues to grow as more early obesity prevention interventions are developed and tested.^16^ Interventions published to date, vary in their effectiveness to reduce childhood obesity and energy-balance related behaviours.^7 16-18^ A potential source of variation in intervention effectiveness may be the components of the interventions.

Intervention components can differ, such as behaviours targeted, delivery features (e.g. mode, setting) and behaviour change techniques (BCTs). Little is known about which components are included in early obesity prevention interventions seeking to change behaviour, and which of those included specific components are and are not effective.^19^ Interventions designed to modify the trajectory of a young child’s growth trajectory are hypothesized to exert their effect by changing parental behaviours that influence children’s energy balance. Traditionally, the different components of behaviour change interventions are under-specified in published reports contributing to a poor understanding of the ways in which effective interventions have their impact (i.e. the ‘black box’ problem).^19^ This limits the ability of researchers and practitioners to optimise, implement and scale up effective interventions that are needed to prevent childhood obesity.^19 20^ Exploring the extent to which the target behaviours, delivery features and BCTs differ between interventions may help to understand why some interventions work better than others.

Deconstructing interventions into their components can provide important information about the parental behaviours that were targeted for change, how an intervention was delivered (i.e. delivery features), and the behaviour change techniques (BCTs; i.e. the smallest, measurable and reproducible behaviour change components)^21^ used to change parents’ behaviour. Deconstructing interventions in this way is possible through the use of ontologies to systematically categorise various intervention components.^21-23^ While there are several reporting checklists, taxonomies and ontologies available to describe behaviour change interventions, the BCT Taxonomy v1 (BCTTv1) to specify BCTs is one of the most commonly used, including examination of obesity-related interventions among adults.^24-26^

Researchers have explored BCTs in parent-focused interventions targeting child obesity-related behaviours, including infant feeding practices, dietary behaviours and physical activity patterns.^27-31^ The proposed work builds on prior work by Martin and colleagues^30^ and Matvienko-Sikar and colleagues^29^ that identified components of interventions targeting obesity, focused on physical activity and eating, and infant feeding interventions (in 2 to 18 year old children and infants, respectively). The current project advances previous reviews by examining interventions commencing antenatally or within one year of birth, covering all obesity-relevant behaviours (relating to infant feeding, dietary intake, physical activity, sedentary behaviour, sleep), drawing on unpublished material describing interventions and using the most comprehensive BCT taxonomy (i.e. BCTTv1).^29 30 32^ To date, no review has comprehensively explored the intervention components of early obesity prevention interventions across multiple behaviours or utilised unpublished intervention materials.

Members of our research team have previously applied a comprehensive approach to better understand factors contributing to the effectiveness of four early obesity prevention interventions undertaken in Australia and New Zealand.^33^ The approach included analysing the content of interventions using descriptions of interventions in both published peer reviewed articles and unpublished materials (e.g. participant manuals, telephone scripts, videos). The number of BCTs identified from published materials only (1 to 11 BCTs per trial)^29^ was much smaller than when including unpublished materials (13 to 25 BCTs per trial),^33^ reinforcing the importance of analysing unpublished intervention materials to obtain a more accurate understanding of such interventions.^34^ This prior work was limited to four trials in one geographical region and results may not be generalisable on a global level. Furthermore, small sample sizes hindered exploration of BCTs by the types of behaviours targeted. We propose extending this innovative approach to include all ongoing and completed trials in this field and analysing BCTs to address all relevant target behaviours in both published and unpublished intervention materials.

The current study will answer the following questions:

1. What are the targeted behaviours, delivery features and behaviour change techniques used in early obesity prevention interventions?
2. What are the similarities and differences in behaviour change techniques used to target different behaviours?
3. Are particular intervention components more effective at reducing obesity risk among children aged around 24 months (i.e. body mass index z-score) than others?

To address these questions, we will code intervention content and evaluate the effectiveness of components to prevent obesity.

## METHODS AND ANALYSIS

This study has been prospectively registered on PROSPERO International prospective register of systematic reviews (CRD42020177408). The current project will complement our individual participant data meta-analysis assessing the effectiveness of early child obesity prevention interventions (Hunter et al. unpublished). A systematic search has been used to identify trials eligible to join the Transforming Obesity in CHILDren (TOPCHILD) Collaboration (www.topchildcollaboration.org), and all eligible trials will be able to contribute to both the current review and the individual participant data meta-analysis. This protocol follows the Preferred Reporting Items for Systematic review and Meta-Analysis protocols (PRISMA) checklist (Supplementary File 1).^35^

### Eligibility criteria

Trials will be included if they 1) are a randomised controlled trial for which randomisation can occur at the individual level or by cluster, including stepped-wedge designs; 2) involve parents/caregivers (including pregnant women) and their infant(s) aged 0 to 12 months at baseline; 3) are evaluating an intervention which continues beyond pregnancy, is child obesity prevention focused and includes at least one behavioural component related to parent feeding practices, early feeding, diet quality, activity/sedentary behaviour or sleep; 4) include a usual care control arm, no intervention or attentional control; 5) include at least one measure of child adiposity measured at the end of intervention (e.g. BMI z-score, prevalence of overweight/obesity, skinfold thickness). Trials will be excluded if they focus solely on obesity in pregnancy, or include non-behavioural interventions (e.g. supplements). See our companion paper for further details (Hunter et al. unpublished).

### Information sources and search strategy

Systematic searches will be conducted annually to identify eligible trials for the duration of the TOPCHILD Collaboration (currently funded until the end of 2023). An initial systematic search was performed on the 18^th^ of March 2020 in the following databases: Medline (Ovid), Embase (Ovid), Cochrane Central Register of Controlled Trials (CENTRAL), CINAHL (EBSCO), PsycINFO. No limits were placed on publication date or language. A search strategy for Medline is presented in Supplementary File 2.

We also searched ClinicalTrials.gov (24^th^ March 2020) and other trial registries via the World Health Organization’s International Clinical Trials Registry Platform (13^th^ May 2020) search portal to identify planned and ongoing trials. Additional trials will be identified by collaborators and contacts notifying the research team of any planned, ongoing or completed trials of which they are aware and will be screened for eligibility.

### Selection process

Two reviewers will independently screen title/abstracts and full text articles against the eligibility criteria, in Covidence systematic review software (Veritas Health Innovation, Melbourne Australia). Agreement between reviewers will be calculated as percent agreement for title/abstract and full text screening. Any disagreements will be resolved by discussion or consulting a third reviewer.

### Data extraction and risk of bias

Eligible trials will be invited by email to nominate a representative/s to join the TOPCHILD Collaboration. Trial representatives (i.e. trialists) will be contacted via email to share unpublished intervention materials for this current review, in English language where possible. Primary analyses will only include trials that have provided both published and unpublished intervention materials, allowing comprehensive intervention coding to be performed. If required, sensitivity analyses will be undertaken to compare intervention components using intervention descriptions reported in published materials of trials that have not shared intervention materials to address potential selection bias. Two reviewers will independently extract general trial characteristics (e.g. author, publication date, intervention name, method of sequence generation and allocation concealment, geographical location, participants) and outcome measures, and record them in FileMaker (FileMaker Pro 18 Advanced; Claris International Inc., Santa Clara, CA, USA). Risk of bias will be assessed for the complementary review examining intervention effectiveness; however, is not required for the current study focused on describing the content of interventions.

### Coding of target behaviours, delivery features and behaviour change techniques

Outcomes for which data will be sought are the discrete intervention components that will be coded by the study team, namely target behaviours, delivery features and BCTs. A standardised procedure will be followed to code intervention materials with a brief training session held to ensure all coders are familiar with the processes to assist consistency in coding target behaviours, delivery features and BCTs. All coders will have completed at minimum the University College London online training for the Behaviour Change Technique Taxonomy v1 (BCTTv1; http://www.bct-taxonomy.com/) and, where possible, have experience in coding BCTs in past projects. All material will be coded by two independent coders, when possible, exceptions may include when unpublished materials are only available in languages other than English. Agreement between coders will be calculated. Any discrepancies in coding will be resolved through discussion; or if no consensus is reached, a third coder will be consulted to reach consensus. The standardised procedure will be used whenever possible, however where unpublished materials are provided in languages other than English a modified procedure will be followed, such as using one coder fluent in the required language resulting in a subset of unpublished materials from a trial being coded once. If necessary, translation services will be sought to ensure the intervention components can be appropriately coded.

**Target behaviours** will be coded to capture the parental behaviour(s) addressed in each intervention. **Table 1** provides examples of behaviours that may be targeted in early obesity prevention interventions. Additional behaviours extracted from trials will be iteratively added to this pre-specified list. Behaviours will be grouped into clusters of behavioural topics, and these may include infant feeding practices, food provision and parent feeding practices, movement practices and sleep health practices (**Table 1**). While eligible interventions can commence antenatally, this study is focused on understanding the behavioural content relating to parental behaviours directed towards infants in the first 12 to 24 months after birth, rather than focusing on parents own health behaviours.

**Table 1:** Examples of specific parental behaviours grouped into clusters of behavioural topics

| Target parental behaviour cluster | Example of specific parental behaviours |
| --- | --- |
| Infant feeding practices | Promoting and/or sustaining breastfeeding, including exclusive breastfeeding to 6 months of age<br>Feeding formula appropriately, if necessary (e.g. making formula per package |
|  | instructions, feeding in response to the infant's hunger/satiety cues, feeling with suitable types of formula)<br>Avoiding unnecessary overfeeding with breastmilk and supplementing with formula<br>Delaying introduction of solid foods (complementary feeding) until 6 months of age |
| Food provision and parent feeding practices | *Behaviours related to dietary intake*<br>Providing appropriate types of foods (e.g. vegetables, meat and alternatives, fruits, whole grains, dairy)<br>Providing age-appropriate portions of each food group (i.e. portion sizes; incl. limiting portions of milk)<br>Limiting provision of certain foods and drinks (e.g. energy-dense, nutrient poor foods, sugar-sweetened beverages)<br>*Behaviours related to feeding practices*<br>Offering foods repeatedly that have previously been rejected<br>Offering foods and drinks in response to infants' hunger/satiety cues (e.g. letting the infant decide how much they eat, not pressuring to eat)<br>Avoiding use of food to control (or reward) the infant's emotions, behaviour or consumption of other foods<br>Providing regular meal routines (incl. eating together, limiting distractions) |
| Movement practices | *Behaviours related to physical activity*<br>Placing infant on their stomach for prone play ('tummy time')<br>Promoting age appropriate physical activity such as active play, outdoor play, activities relating to fundamental movement skills<br>Providing toys that promote movement such as balls and toys on wheels<br>*Behaviours related to sedentary behaviour*<br>Limiting the amount of time the infant is restrained (e.g. prams/strollers, high chairs, strapped on a caregivers back)<br>Limiting the amount of time the infant is exposed to screens (e.g. television, mobile devices)<br>Providing alternatives to screen time |
| Sleep health practices | Promoting regular sleep routine (e.g. calm, quiet, soothing)<br>Letting the infant settle back to sleep when stirring/crying during sleep cycle (e.g. leaving the room, only picking up infant when awake)<br>Promoting a positive sleep environment (e.g. quiet, darkened, warm)<br>Placing infant in cot/bassinet while awake and letting infant learn to fall asleep (e.g. following infant's signs of tiredness)<br>Avoiding bed-sharing / co-sleeping (i.e. sleeping with the infant in the same bed)<br>Maximising day-night differences (e.g. lights on and play in the day, lights off and sleep at night) |

**Delivery features** refer to a broad number of intervention characteristics that relate to how an intervention is delivered. Delivery features will include items in the Template for Intervention Description and Replication (TIDieR) reporting checklist,^36^ such as who conducted the intervention, how (mode of delivery), where (setting), when and how much (intensity), how well the intervention was delivered (fidelity), and if there were modifications made at the intervention level (**Table 2**). Draft ontologies from the Human Behaviour Change Project^22^ will be used to code the intervention setting (Intervention Setting Ontology), mode of delivery (Mode of Delivery Ontology) and source delivering the intervention (Intervention Source Ontology). Such ontologies provide a common language to describe and compare several delivery features across interventions. Delivery features that cannot be classified using existing checklists/ontologies will be added as additional categories.

**Table 2:** Delivery features and corresponding Human Behaviour Change Project ontologies and project-developed categories based on the TIDieR framework

| **Delivery features<sup>1</sup>** | **Example categories** |
| --- | --- |
| **Why – theory:** Rational, theory or goal | *Theory name and / or factors identified as needing to change reported in intervention* |
| **What – materials:** Physical or informational materials, including provided to participants | DVD / video<br>Written materials<br>Newsletters<br>PowerPoint slides<br>Website<br>Mobile application |
| **What – procedures:** Procedures, activities, processes used in the intervention | Didactic sessions<br>Group discussion<br>Peer support |
| **Who provided – intervention delivered by:** Expertise, background and any specific training (for each intervention provider) | *Intervention Source Ontology*<br>e.g.<br>Nursing professional<br>Community health worker<br>Dietician and Nutritionist |
| **How – delivery mode:** (includes delivery to individuals or groups) | *Mode of Delivery Ontology*<br>e.g.<br>Face to face<br>Letter<br>Mobile digital device |
| **Where – intervention setting:** Location | *Intervention Setting Ontology*<br>e.g.<br>Household residence<br>Community healthcare facility |
| **When and how much – intervention dose:** | Total number of contacts<br>Frequency of contact: < weekly, weekly to <monthly; monthly or greater<br>Duration of contact: brief, moderate, extended |
| **Tailoring:**<br>If the intervention was planned to be personalised, titrated or adapted at the participant level | Yes – there was an element of tailoring in the intervention<br>No<br>Not described |
| **Modifications:**<br>If the intervention was modified during the study at the intervention level | Yes – the intervention was modified<br>No<br>Not described |
| **Fidelity:**<br>Planned and Actual | *Fidelity of the intervention extracted as reported in the intervention* |

TiDieR: Template for Intervention Description and Replication
<sup>1</sup> Adapted from Hoffmann et al.<sup>36</sup>

**Behaviour Change Techniques** will be coded using the BCTTv1.^21^ This taxonomy was developed through a consensus process with experts from a range of disciplines from several countries, and selected for the current study as a multidisciplinary standardised language to categorise intervention content.^21^ Standard coding procedures will be followed, for example the whole intervention description will be read before coding.^2^ Behaviour Change Techniques that are clearly present in the intervention from the description provided will be coded as ‘Yes’, and BCTs that are likely present but with insufficient evidence will be coded as ‘Maybe’.^37 38^ To be coded as ‘Yes’ the BCTs are required to target parents (i.e. target population) and a parental behaviour related to the target behavioural clusters as described in Table 1. Due to the complex number of different target behaviours across eligible trials and the scope of this project, BCTs will be coded to the target behaviour cluster rather than each individual target behaviour. Each BCT identified will be coded to the relevant target behaviour cluster/s when there is sufficient detail to separate content in this way. When this is not possible BCTs will be coded to ‘unspecified behavioural cluster’. Intervention content will be coded from both published (e.g. protocols, main results, and follow-up publications) and unpublished intervention materials (e.g. participant manuals, telephone scripts, videos). Access to unpublished materials is important to understand details of an intervention and allow coding of additional BCTs not reported in published descriptions.^19 34^ Control arms will also be coded for the presence of BCTs relevant to the target population and behaviours, and only BCTs unique to the intervention arm will be used in the results synthesis.^20^ Two trained coders will perform and record coding in Microsoft Excel. Agreement of initial coding between coders will be calculated by kappa and prevalence-adjusted bias-adjusted kappa (PABAK) statistics to assess strength of agreement.^39^

Following agreement between coders, a validation process will be undertaken. Coded BCTs for each target behaviour cluster from published and unpublished materials for each trial will be sent to the respective trialists to validate the coding. Where possible, a virtual meeting will be organised for the coder to discuss the coding with the trial representative and to minimize reliance on trialists knowledge of BCTs. If there are discrepancies between the coder and trial representative, these will be discussed to reach consensus, including referring to the intervention materials as the primary source of evidence.

### Synthesis of results

To address the first research question, a structured summary will be prepared to describe the targeted behavioural clusters, delivery features and BCTs used in early obesity prevention interventions. To address the second research question, narrative comparisons of BCTs used by target behaviour clusters will be made to explore the similarities and differences in BCTs used to target different behaviours. To address the third research question, exploratory analyses will be undertaken to provide preliminary information about the effectiveness of commonly used intervention components in reducing body mass index (BMI) z-scores at 2 years of age (+/-6 months). For this purpose, a meta-regression analysis will be performed for each commonly used intervention component (i.e. used in 5 or more interventions), to compare infant BMI z-score for trials including the intervention component compared to trials not including the intervention component. Our proposed approach will take into account small sample sizes and importantly the variability of the observed effect sizes, however, will not be able to determine independent effects of each component.

### Patients and public involvement

The TOPCHILD Collaboration involves a broad range of stakeholders including health professionals, policy makers, researchers and trialists. In addition, the Collaboration includes a parent representative and an intervention facilitator/nurse who have given input into and feedback on this protocol and will be involved in the interpretation of results.

## ETHICS AND DISSEMINATION

The study has been approved by The University of Sydney Human Research Ethics Committee (project no. 2020/273) and Flinders University Social and Behavioural Research Ethics Committee (project no. HREC CIA2133-1). If any amendments to this published protocol are required, they will be documented in the PROSPERO registration record (no. CRD42020177408).

Findings from the current study will be disseminated through peer-reviewed publications, conference presentations, and targeted communication with key stakeholders, such as intervention designers. Disseminating findings to intervention designers will impart knowledge about common intervention approaches used in the field of early obesity prevention as well as less commonly used but potentially effective BCTs and delivery features that can be explored in future interventions.

## DISCUSSION

Our study will characterise infant obesity prevention interventions commencing antenatally or in the first 12 months after birth, by specifying the targeted behaviours, delivery features and applied BCTs. Key strengths of this study include the comprehensive systematic search to identify planned, ongoing, and completed early childhood obesity prevention trials that will provide a broad understanding of the behaviour change content and delivery features used around the world. By looking into the ‘black box’ of interventions, this study will provide detailed summaries of methodologies used in early childhood obesity prevention interventions globally. We are extending previous methods by coding BCTs in both unpublished and published materials and performing cross validation of coding with trialists through the TOPCHILD Collaboration. In addition, we are exploring patterns in BCT use across four key obesity prevention parental behaviour clusters; namely infant feeding practices, food provision and parent feeding practices, movement practices, and sleep health practices. We will use standardised coding taxonomies (i.e. BCTTv1), and pilot test new ontologies from the Human Behaviour Change Project^22^ to systematically code intervention source, mode of delivery and intervention setting. This study will provide preliminary insights into which intervention components are more effective than others. However, because BCTs are not used in isolation and interventions include multiple components, it is not possible to isolate the individual effects of each BCT or component within a trial or across trials from the effects of other BCTs, and there may be confounding through unobserved trial-level effects. Intervention coding will be limited to indicating the presence or absence of a BCT in intervention materials. Coding will not address whether techniques were in fact delivered to each participant (i.e. fidelity of BCT) or BCT dose. Nevertheless, we hope that this exploratory analysis will provide preliminary insight into which intervention components may be more effective than others.

A systematic understanding of the components of early obesity prevention interventions will lay the groundwork for conducting quantitative predictive modelling in future research projects. The current study will generate a comprehensive database of intervention components for each trial in the TOPCHILD Collaboration in standardised terminology and classified by target behaviours, delivery features and BCTs. The resulting database will be combined with individual participant data obtained from TOPCHILD trialists (see Hunter et al, unpublished protocol) in a future study to perform quantitative predictive modelling. Predictive modelling will further our understanding of effective intervention components for reducing childhood obesity, including identification of components that are particularly effective for key population groups. Project updates will be publicly available on the TOPCHILD Collaboration website at https://www.topchildcollaboration.org/.

## Data Availability

No data are available for the current manuscript since this is a protocol.

## Acknowledgements

We would like to acknowledge Slavica Berber from the NHMRC Clinical Trials Centre for advice on the search strategy. We would also like to acknowledge the NHMRC Centre of Research Excellence in Early Prevention of Obesity in Childhood, who supported the pilot and foundational work for this project.

We would like to thank Maria Luisa Garmendia and Camila Corvalan for their feedback on an earlier version of this protocol.

## TOPCHILD Collaboration members

### Steering Group

Anna Lene Seidler, Kylie Hunter, Brittany Johnson, Rebecca Golley, Lisa Askie, Angie Barba, Mason Aberoumand, Sol Libesman

### Advisory Group

Alison Hayes, Charles Wood, Chris Rissel, David Espinoza, Denise O’Connor, Ian Marschner, Kristy Robledo, Louise Baur, Lukas Staub, Luke Wolfenden, Michelle Sue-See, Paul Chadwick, Rachael Taylor, Sarah Taki, Seema Mihrshahi, Wendy Smith, Shonna Yin, Lee Sanders

### Trial Representatives (to date)

Alison Karasz, Amanda Thompson, Ana Maria Linares, Ana Perez Exposito, Ata Ghaderi, Barry Taylor, Carolina González Acero, Cathleen Odar Stough, Claudio Maffeis, Cristina Palacios, Christine Helle, Eliana Perrin, Emily Oken, Eva Corpeleijn, Finn Rasmussen, Heather Wasser, Hein Raat, Ian Paul, Jennifer Savage, Jessica Thomson, Jinan Banna, Junilla Larsen, Karen Campbell, Kaumudi Joshipura, Kayla de la Haye, Ken Ong, Kylie Hesketh, Levie Karssen, Li Ming Wen, Lynne Daniels, Margrethe Røed, Maria Bryant, Maribel Campos Rivera, Mary Jo Messito, Michael Goran, Nina øverby, Rachael Taylor, Rachel Gross, Rajalakshmi Lakshman, Russell Rothman, Sarah-Jeanne Salvy, Sharleen O’Reilly, Stephanie Anzman-Frasca, Vera Verbestel

## Authors’ contributions

ALS together with KEH, BJJ, LA, RKG conceived the idea for the study.

BJJ, KEH, ALS, RKG, LA developed the research question and protocol registration.

BJJ wrote the first draft of the manuscript.

KEH, BJJ, ALS, RKG, LA developed the eligibility criteria and KH developed the search strategy.

KEH, MA, AB, BJJ, SL, ALS performed the search and screening.

BJJ, PC, RKG developed the coding procedure.

AJH, CTW, CR, DE, DAO’C, ICM, KPR, LAB, LPS, LW, MS-S, PC, RWT, ST, SM, WS provided critical review and feedback at each stage of the process.

All authors critically revised the manuscript for intellectual content, and agreed and approved the final manuscript. BJJ is the guarantor of the manuscript.

