## Supplementary file 2 for "Unpacking the behavioural components and delivery features of early childhood obesity prevention interventions in the TOPCHILD Collaboration: a systematic review and intervention coding protocol"

Supplementary file 2: Example of the TOPCHILD Collaboration search strategy

**Medline Search Strategy:**

**Ovid MEDLINE(R) ALL 1946 to March 16, 2020**

1. pediatric obesity/
2. Weight Gain/
3. obes*.ti,ab
4. (weight gain).ti,ab
5. (overweight or over weight).ti,ab
6. weight change*.ti,ab
7. ((bmi or body mass index) adj2 (gain or loss or change)).ti,ab
8. 1 or 2 or 3 or 4 or 5 or 6 or 7
9. social support/
10. ((behaviour or behavior) and change).ti,ab
11. ((behavio?r*) adj (therapy or modif* or strateg* or intervention* or advice or program* or class* or counsel* or educat* or instruct* or teach* or train* or guidance or lesson* or workshop* or module* or consultation* or session*)).ti,ab
12. ((lifestyle or life style) adj (chang* or modif* or strateg* or intervention* or advice or program* or class* or counsel* or educat* or instruct* or teach* or train* or guidance or lesson* or workshop* or module* or consultation* or session*)).ti,ab
13. social support.ti,ab
14. (peer adj2 support).ti,ab
15. counsel?ing.ti,ab
16. education* adj1 (intervention* or program* or class* or counsel* or teach* or workshop* or module* or consultation* or session*)).ti,ab
17. home visit*.ti,ab
18. 9 or 10 or 11 or 12 or 13 or 14 or 15 or 16 or 17
19. exp Breastfeeding/
20. Infant Nutritional Physiological Phenomena/
21. Child Nutrition Sciences/
22. Infant Food/
23. ((child or toddler or infant$) adj1 (food or feeding or nutrition$)).tw.
24. ((responsive or complementary) adj1 feeding).ti,ab
25. ((diet* or nutrition) adj (modif* or strateg* or intervention* or advice or program* or class* or counsel* or educat* or instruct* or teach* or train* or guidance or lesson* or workshop* or module* or consultation* or session*)).ti,ab
26. (healthy eating).ti,ab
27. (fruit or vegetable*).ti,ab
28. (high fat* or low fat* or fatty food*).ti,ab
29. 19 or 20 or 21 or 22 or 23 or 24 or 25 or 26 or 27 or 28
30. exp Exercise/
31. exercis*.ti,ab
32. (physical activity or physical inactivity).ti,ab
33. sedentary behavio?r.ti,ab
34. (screen time).ti,ab
35. 30 or 31 or 32 or 33 or 34
36. Sleep/
37. Exp Primary prevention
38. exp Health Promotion/
39. exp Health Education/
40. prevention.mp
41. prevent*.ti,ab
42. (health promotion or health education or health communication).ti,ab
43. exp Obesity/pc (Prevention and Control)
44. exp Overweight/pc (Prevention & Control)
45. (obesity adj2 prevent*).ti,ab
46. (overweight adj2 prevent*).ti,ab
47. 37 or 38 or 39 or 40 or 41 or 42 or 43 or 44 or 45 or 46
48. 8 and (18 or 29 or 35 or 36) and 47
49. exp child/ or exp infant/
50. ((child* or infant* or baby or toddler* or pediatr* or paediatr*) not adolescen*).ti,ab
51. (pregnan* or antenatal or parent or parent$1 or care giver or caregiver or guardian or family or families or mother$1 or father$1).ti,ab
52. 49 or 50 or 51
53. 48 and 52
54. (exp animals/ not humans.sh.) or (rat or rats or mouse or mice or rodent*).ti.
55. 53 not 54
56. controlled clinical trial.pt.
57. randomi#ed.ti,ab.
58. randomly.ab.
59. (clinical trials as topic or controlled clinical trials as topic).sh.
60. trial.ti.
61. exp randomized controlled trial/ or exp randomized controlled trials as topic/
62. 56 or 57 or 58 or 59 or 60 or 61
63. 55 and 62

### WHO ICTRP

**Search string**

Basic search

1. infant AND obesity prevention

2. infant AND prevention of obesity

3. infant AND overweight prevention

4. infant AND prevention of overweight

5. infant AND prevent AND obesity

6. child AND obesity prevention

7. child AND prevention of obesity

8. child AND overweight prevention

9. child AND prevention of overweight

10. child AND prevent AND obesity

Advanced search

1. Title: prevent AND obesity

Recruitment Status: All

Limit: Search for clinical trials in children

2. Title: prevent AND overweight

Recruitment Status: All

Limit: Search for clinical trials in children

3. Title: prevent OR prevention

Condition: obesity OR overweight

Recruitment Status: All

Limit: Search for clinical trials in children
